# Efficacy of LD Bio *Aspergill*us ICT Lateral flow assay for serodiagnosis of chronic pulmonary aspergillosis

**DOI:** 10.1101/2022.03.04.22271825

**Authors:** Animesh Ray, Mohit Chowdhury, Janya Sachdev, Prayas Sethi, Ved Prakash Meena, Gagandeep Singh, Immaculata Xess, Surabhi Vyas, MA Khan, Sanjeev Sinha, David W. Denning, Naveet Wig, SK Kabra

## Abstract

**Background:** The diagnosis of CPA relies on the detection of IgG *Aspergillus* antibody which is not freely available, especially in resource-poor settings. Point-of-care tests like LDBio *Aspergillus* ICT lateral flow assay, evaluated in only a few studies, have shown promising results for diagnosis of CPA. However no study has compared the diagnostic performances of LDBio LFA in setting of tuberculosis endemic countries and have compared it with that of IgG *Aspergillus*.

**Objectives:** This study aimed to evaluate the diagnostic performances of LDBio LFA in CPA and compare it with existing diagnostic algorithm utilising ImmunoCAP IgG Aspergillus.

**Methods:** Serial patients presenting with respiratory symptoms (cough, hemoptysis, fever etc) for > 4 weeks were screened for eligibility. Relevant investigations including direct microscopy and culture of respiratory secretions, IgG *Aspergillus*, chest imaging etc were done according to existing algorithm. Serum of all patients were tested by LDBio LFA and IgG *Aspergillus* (ImmunoCAP Asp IgG) and their diagnostic performances were compared.

**Results:** A total of 174 patients were included in the study with ∼66.7% patients having past history of tuberculosis. A diagnosis of CPA was made in 74 (42.5%) of patients. The estimated sensitivity and specificity of LDBio LFA was 67.6% (95% CI: 55.7%-78%) and 81% (95% CI: 71.9%-81%) respectively which increased to 73.3% (95% CI: 60.3%-83.9%) and 83.9% (95% CI: 71.7%-92.4%) respectively in patients with past history of tuberculosis. The sensitivity and specificity of IgG *Aspergillus* was 82.4% (95% CI: 71.8%-90.3%) and 82% (95% CI: 73.1-89%); 86.7% (95% CI: 75.4%-94.1%) and 80.4% (95% CI: 67.6%-89.8%) in the whole group and those with past history of tuberculosis respectively.

**Conclusions:** LDBio LFA is a point-of-care test with reasonable sensitivity and specificity. However, further tests may have to be done to rule-in or rule-out the diagnosis of CPA in the appropriate setting.

## Introduction

The relationship between the fungus and the host typically determines the manifestation of *Aspergillus* lung disease ranging from acute and subacute invasive to chronic pulmonary aspergillosis.^1^,^2^ Chronic pulmonary aspergillosis (CPA) is a collection of serious illnesses characterised by persistent cough, dyspnea, haemoptysis, fatigue and weight loss.^3^ Chronic cavitary pulmonary aspergillosis (CCPA) is the most frequent manifestation of CPA, which can progress to chronic fibrosing pulmonary aspergillosis if left untreated. Single aspergilloma and *Aspergillus* nodule are some of the less prevalent manifestations of CPA.^4^ Pre-disposing factors for CPA include underlying pulmonary illnesses like chronic obstructive pulmonary disease (COPD) or mycobacteriosis, as well as prevalent immunosuppressive disorders like diabetes.^3^

Patients with CPA have significant morbidity affecting roughly 3 million individuals globally. The overall 5-year mortality rate ranges up to 80%, resulting in an estimated 450,000 yearly fatalities.^5^ As a result of India’s high TB disease load, post-tuberculosis sequelae are common.^6^ CPA has an annual incidence of 27,000 to 170,000 cases, with a 5-year prevalence of 24 per 100,000.^7^ In India, the prevalence may be very high in post-tuberculosis sequelae patients (∼57%) and recurrence of tuberculosis has been reported to be an important independent risk factor for development of CPA. ^8^ No single clinical or radiological manifestation or laboratory result is adequate for a conclusive diagnosis of CPA; rather, an amalgamation of clinical, radiographic, and microbiological findings is used since the presentation may be non-specific and might be difficult to differentiate from pulmonary tuberculosis.^9^,^10^,^11^,^12^ The microbiological evidence for diagnosis is considered with direct confirmation of *Aspergillus* infection (microscopy or culture from BAL fluid/biopsy) or a immune response to *Aspergillus* spp.^4^

In the frequent absence of positive cultures, serologic assays are essential for diagnosis of CPA.^13^ In the earliest assay formats, antibodies against *Aspergillus fumigatus* were identified by detection of precipitins with high specificity utilising the double immunodiffusion test (DID) or counter immune-electrophoresis (CIE) technique. These methods however had a long turnaround time, required significant labour with a large inoculum of fungal extract and patient serum extracts, and the results were only semi-quantitative.^13^ Other commercially accessible serological tests subsequently were launched which may be employed for diagnosis of CPA, such as enzyme immunoassay (EIA), enzyme-linked immunosorbent assay (ELISA) and indirect hemagglutination (IHA), however performance levels vary amongst tests, and redefining of cut-off values for different populations and diagnoses may be required to maximise performance.^9^ Indirect hemagglutination is clearly inferior to other methods.^14 14^Moreover, performance of these tests often requires costly equipment, steady power supply, technical expertise as well as considerable costs.

In recent years, the lateral flow assay (LFA) has been employed to simplify Aspergillus IgG detection with quick turn-around time and little laboratory equipment. The only commercially available LFA for detecting *Aspergillus* IgG is LDBio LFA Aspergillus immunochromatographic technology (hereafter referred to as LDBio LFA).^15^ A point-of-care test has been felt to be essential in simplifying the diagnosis and management of CPA, especially in a resource-limited setting. When compared to ImmunoCAP (Thermo Fisher) for levels of Aspergillus fumigatus specific IgG (hereafter referred to as ImmunoCAP Asp IgG), the LDBio LFA has exhibited good sensitivity and specificity for detection of CPA in studies done in centres in France and United Kingdom.^16, 17^ A study had also reported sensitivity and specificity of 80% and 70% respectively of LDBio LFA from Indonesia, which however did not use an alternative IgG *Aspergillus* in the diagnostic algorithm. So, the present study was conducted out of the necessity of evaluating the diagnostic performances of this point-of-care test in a tuberculosis endemic country with a significant burden of CPA and to compare it with the existing diagnostic criteria including IgG *Aspergillus*.

## Materials & Methods

Between February 2020 and December 2021, consecutive patients presenting to chest clinic of a tertiary care unit in North India with respiratory symptoms (cough, hemoptysis, fever, shortness of breath, chest pain etc) of more than four weeks duration were enrolled in the study. Patients with an apparent non-CPA diagnosis such as lung malignancy or who refused consent for serological tests were excluded from the study. Ethical permission was taken from Institute Ethics Committee for the conduct of this study (IEC-52/08.01.2021, RP-10/2021). Written consent form was obtained from all participants before enrolment into the study as per institutional protocol.

After enrolment, the case details were recorded in a standardized case record form (CRF), by a trained professional. Relevant investigations including blood tests, chest imaging, sputum examination, bronchoalveolar lavage was done as per the discretion of the treating physician. Serum samples collected from all the individuals were evaluated by LDBio LFA assays and ImmunoCAP Asp IgG assay. The diagnosis of CPA was made individually by two researchers (AR and MC), and then corroborated as per the European Respiratory Society/European Society of Clinical Microbiology and Infectious Diseases (ERS/ESCMID) criteria.^18^ The diagnosis relied on appropriate clinical, radiological and microbiological parameters. Microbiological evidence included a positive serological result using the ImmunoCAP Asp IgG assay to measure *Aspergillus*-specific IgG levels (> 27 mgA milligrams of antibodies]/liter considered to be the cut-off for positive result), histopathological evidence of CPA following lung biopsy or resection, positive result in galactomannan assay performed on serum or BAL samples using the Platelia Aspergillus galactomannan ELISA (Bio-Rad Laboratories) interpreted according to cutoffs provided in the 2019 EORTC/MSGERC guidelines (galactomannan index > 1 was considered positive for both serum and BAL),and respiratory samples showing hyaline septate hyphae morphologically suggestive of *Aspergillus* spp in direct microscopy, or growth of *Aspergillus* spp in culture^4^. Radiological criteria for diagnosis were adapted from those used commonly in a resource-constrained setting ^19^ The *Aspergillus* ICT IgG IgM lateral flow assay (LDBio LFA, Diagnostics, Lyon, France) was used to test each sample, and all tests were performed and interpreted according to the manufacturer’s instructions. The presence of a well-defined black line at the “Test (T)” and “Control (C)” markers was considered as a positive result. The presence of a thin, diffuse grey line at the “T” marker indicates a “weakly positive” result..

An alternative analyses was done assuming cut-off of 40mgA/L for IgG against Aspergillus spp, as is suggested by the manufacturer as well as validated in studies.^17 20 21^

### Statistical analysis

The sample size was estimated considering a sensitivity of 85% and specificity of 84% (average of the diagnostic accuracies reported by Hunter et al ^17^ in healthy population and Rozaliyani et al^22^in tuberculosis-treated patients) and taking a precision of 8.5% with 95% confidence level. The sample size estimated was at least 68 cases and 72 non-CPA cases.

Categorical and continuous variables were reported in frequencies and in percentages and mean with standard deviation/median with minimum, maximum depending on the nature of the data respectively. Fisher’s exact /chi-squared tests were used to establish association for categorical variables while Student’s t-test or Wilcoxon ranksum test was used for continuous variables as appropriate. Those with p value < 0.05 were considered to be significant.

## Results

A total of 218 patients were screened and 174 patients were included in this study (figure 1). The baseline characteristics of the study population are detailed in Tables 1-3. One hundred and sixteen (62.1%) enrollees were male with a mean age of 40.7 (±13.9 years). The vast majority of patients (98.3%) were HIV seronegative. In total, 116 (66.67%) patients had a prior history of documented pulmonary tuberculosis and 46 (29.1%) had used inhalational devices in the last month.

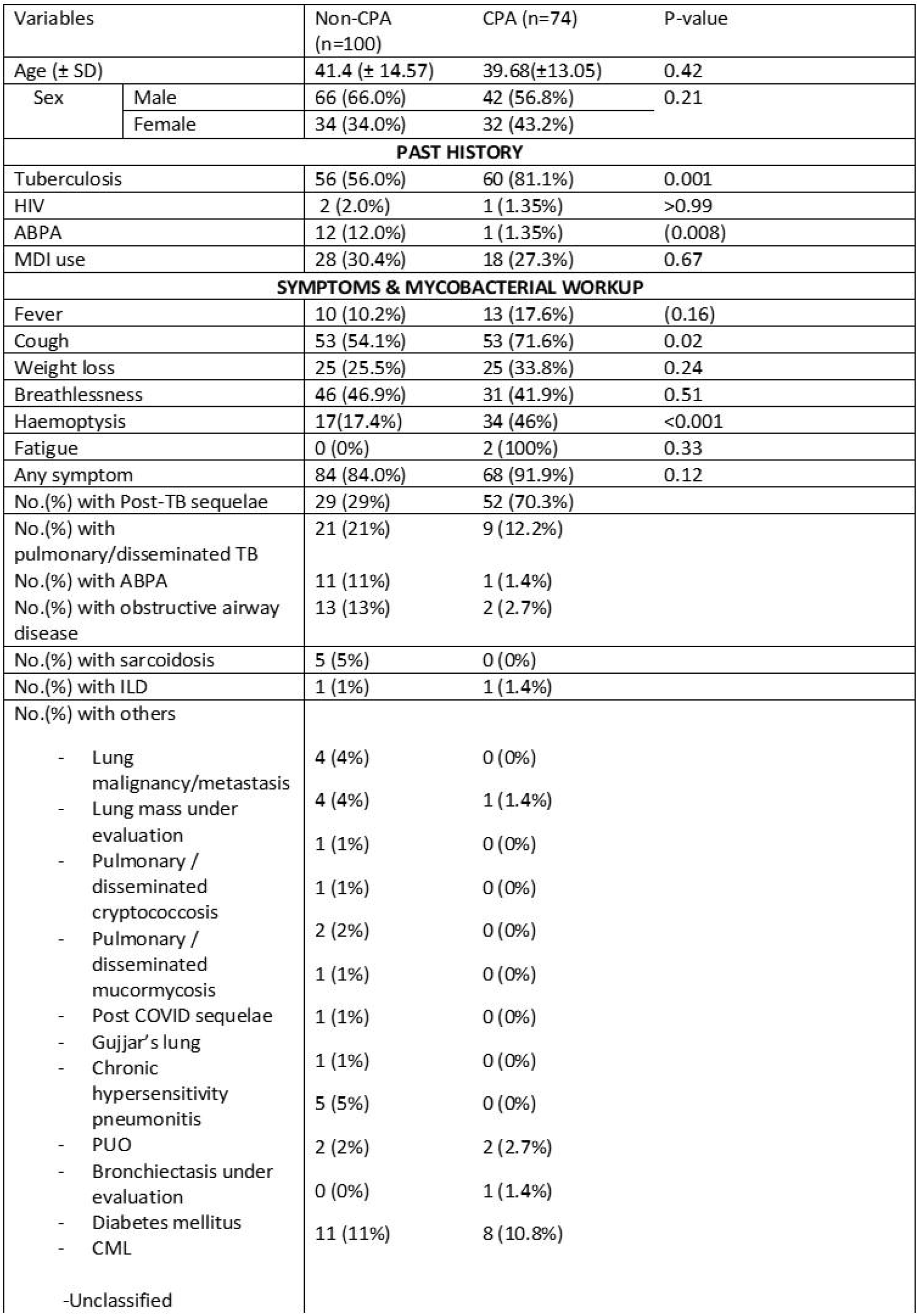

| Features | Non-CPA (n=100) | CPA (n=74) | P-value |
| --- | --- | --- | --- |
| Consolidation | 33 (39.8%) | 35 (48.6%) | 0.33 |
| Cavity | 22 (29.3%) | 61 (85.9%) | <0.001 |
| Nodules | 40 (49.4%) | 42 (61.8%) | 0.14 |
| Ground glass opacities | 22 (30.1%) | 26 (41.3%) | 0.21 |
| Pleural effusion | 16 (22.2%) | 5 (8.8%) | 0.06 |
| Pleural thickening | 10 (10.3%) | 24 (32.9%) | <0.001 |
| Bronchiectasis | 36 (36%) | 46 (62.2%) | 0.001 |

| Variables | Non-CPA (n=100) | CPA (n=74) | P-value |
| --- | --- | --- | --- |
| LDBio LFA positive | 19 (19%) | 50 (67.6%) | <0.001 |
| *Weakly positive* | 2 (2%) | 1 (1.4%) | 0.6 |
| Specific IgE (IQR) | 0.11 (0.02-0.37) | 0.34 (0.04-1.5) | 0.2 |
| Total IgE (IQR) | 328 (66.9-1465) | 406 (119-1035) | 0.8 |
| Specific IgG (IQR) | 11.5 (6.40-20.1) | 53.85 (30-91) | <0.001 |
| AEC (IQR) | 162.07 (49.6-341.66) | 162.32 (15.19-371.46) | 0.93 |
| Specific IgE (≥ 0.1 KVA/L) | 16 (16/28, 57.1%) | 30 (30/46, 65.2%) | 0.49 |
| Total IgE (≥ 500 KVA/L) | 14 (14/31, 45.2%) | 20 (20/47, 42.6%) | 0.82 |
| Specific IgG (≥ 27 MgA/L) | 18 (18.0%) | 61 (82.4%) | <0.001 |
| AEC (≥ 500 cells/mm3) | 13 (13/60, 21.7%) | 10 (10/50, 20%) | 0.83 |
| Positive direct KOH | 4 (4/55, 7.8%) | 6 (6/53, 11.3%) | 0.52 |
| Positive fungal culture | 2 (2/70, 2.9%) | 7 (7/61, 11.5%) | 0.08 |
| *Aspergillus fumigatus* |  | 2 |  |
| *Aspergillus flavus* | 2* | 3 |  |
| *Aspergillus niger* | 1* | 1 |  |
| *Aspergillus* spp. |  | 1 |  |
| Serum galactomannan (≥1.0) | 10 (10/28, 35.7%) | 16 (16/39, 41%) | 0.66 |
| BAL galactomannan (≥1.0) | 7 (7/30, 23.3%) | 17 (17/28, 60.7%) | 0.004 |
| ZN AFB +ve | 2 (2/48, 4.2%) | 0 (0/41, 0%) | 0.49 |
| MGIT +ve | 5 (5/30, 16.7%) | 0 (0/25, 0%) | 0.06 |
| GeneXpert | 9 (9/55, 16.4%) | 4 (4/54, 7.4%) | 0.24 |
| Any TB investigation +ve | 12 (12/100, 12.0%) | 4 (4/74, 5.4%) | 0.19 |

\*One patient showed mixed growth of *Aspergillus niger* and *Aspergillus flavus*
IQR: Interquartile range
AEC: absolute eosinophil count
ZN AFB: ZIEHL-NEELSEN acid-fast bacillus
MGIT: Mycobacteria Growth Indicator Tube

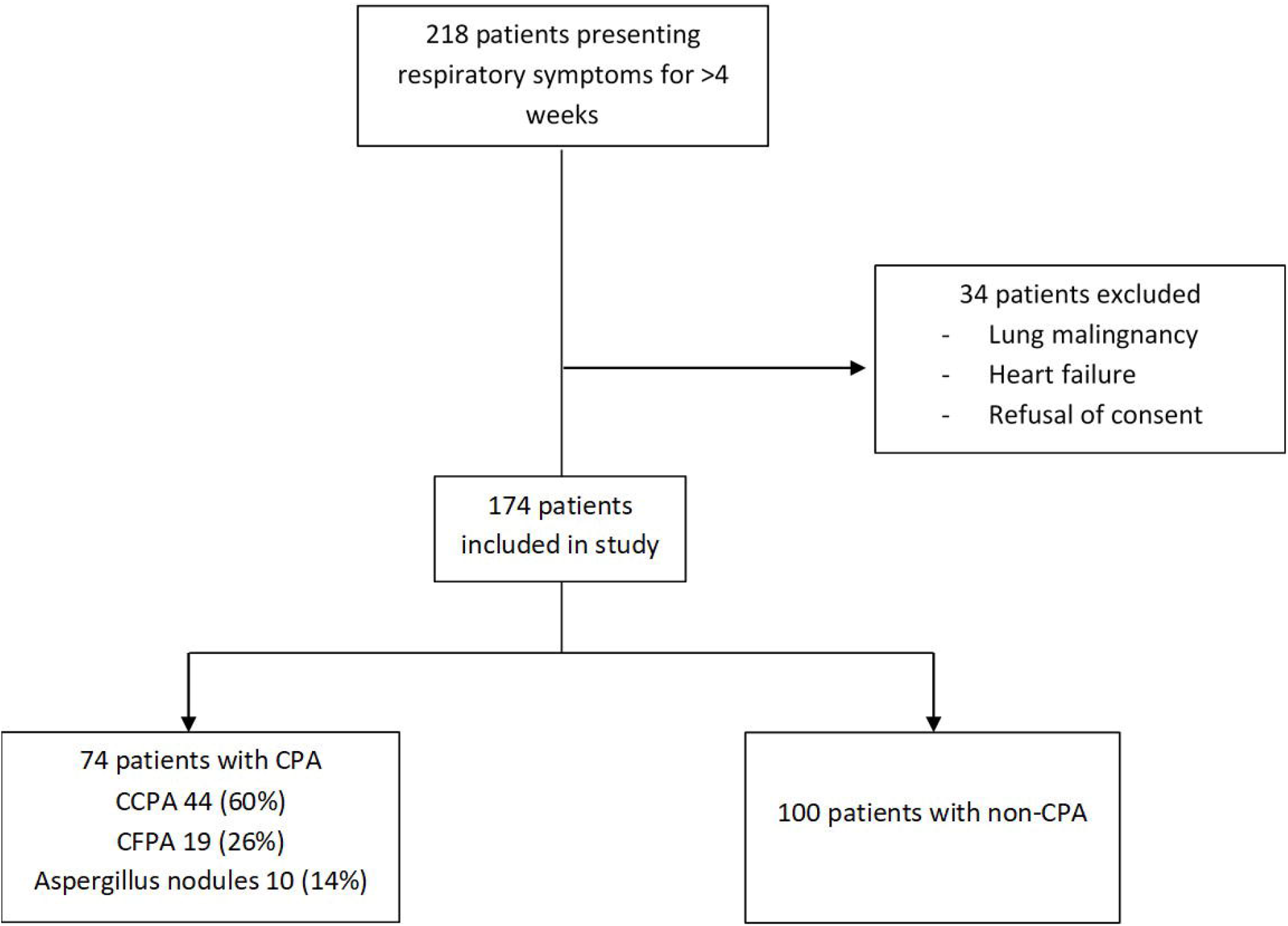

In this study population, 74 patients (42.5%) fulfilled the diagnosis of CPA while a diagnosis of allergic bronchopulmonary aspergillosis was made in 13 (7.5%) patients. Out of the 74 patients with CPA 44 (60.3%) had chronic cavitary pulmonary aspergillosis, 19 (26%) chronic fibrosing pulmonary aspergillosis and 10 (13.7%) had *Aspergillus* nodules. Majority of the patients(> 90%) was diagnosed by positive ImmunoCAP Asp IgG assay as the sole microbiological criteria along with corroborative clinic-radiological features. Pulmonary tuberculosis was diagnosed in 16 (9.2%) patients. Cough was the most common symptom, reported in 106 (61.6%) patients. On chest imaging, cavity was the most common reported finding, being present in 74 patients (42.5%). Serum IgG specific for *Aspergillus fumigatus* was elevated in 61 (82.4%) of the group diagnosed with CPA. Out of 131 patients who produced sputum or underwent bronchoalveolar lavage (BAL), fungal culture showed growth of *Aspergillus* spp in nine patients (6.9%). BAL galactomannan was positive in 17 (9.8%) patients in those with CPA.

The sensitivity and specificity of LDBio LFA for diagnosis of CPA (as compared to the ERS/ESCMID criteria) in our study subjects presenting with respiratory symptoms for at least four weeks was 67.6% (95% CI: 55.7%-78%) and 81% (95% CI: 71.9%-81%) respectively with a diagnostic accuracy of 75.3%. In the population with a past history of tuberculosis, the sensitivity and specificity improved to 73.3% (95% CI: 60.3%-83.9%) and 83.9% (95% CI: 71.7%-92.4%) respectively; and the estimated diagnostic accuracy was 78.5%. In those with past history of tuberculosis and with symptoms >3 months, the sensitivity and specificity further improved to 74.1% (95% CI: 60.3%-85%) and 85% (95% CI: 70.2%-94.3%) respectively; with diagnostic accuracy being 78.7%.

In contrast, the sensitivity and specificity for ImmunoCAP Asp IgG for diagnosis of CPA was 82.4% (95% CI: 71.8%-90.3%) and 82% (95% CI: 73.1-89%) respectively. The diagnostic performances of both LDBio LFA and IgG are represented in Table 4. The kappa for agreement between LDBio LFA and *Aspergillus fumigatus* was 0.53.

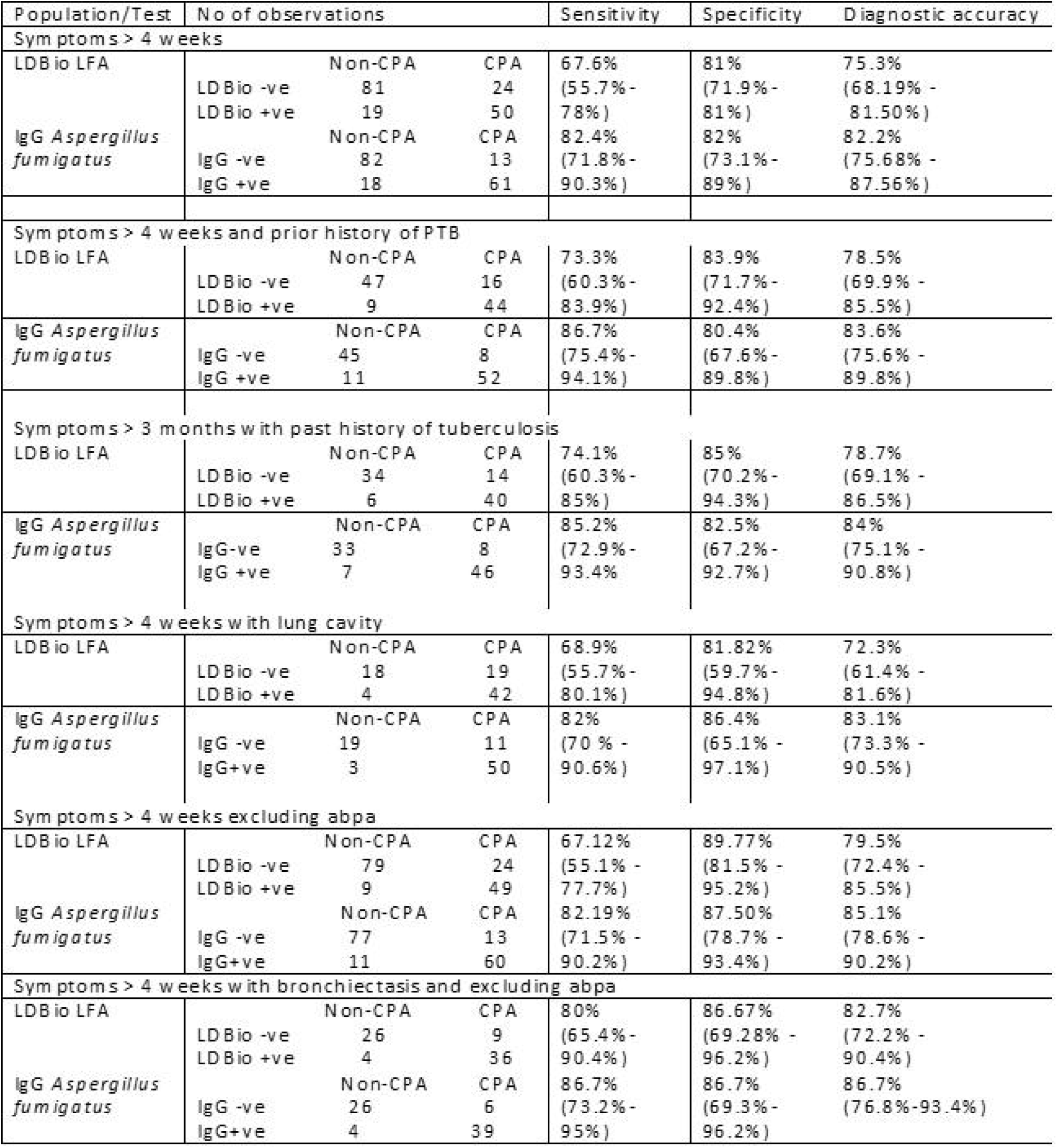

According to the alternative analyses using cut-off of Aspergillus-specific IgG assay as 40 mgA/L, the diagnostic performances of LDBio LFA were altered (in most cases the diagnostic accuracies marginally fell) as shown in Table 5. Chest radiograph and CT thorax of a CPA patient who was negative for both ImmunoCAP Asp IgG & LDBio LFA is depicted in figure 3

| Population/Test No of observations |  |  |  | Sensitivity | Specificity | Diagnostic accuracy |
| --- | --- | --- | --- | --- | --- | --- |
| Symptoms > 4 weeks |  |  |  |  |  |  |
| LDBio LFA |  | Non-CPA | CPA | 68.3% | 75.4% | 72.99% |
|  | LDBio -ve | 86 | 19 | (55.04% - | (66.49% - | (65.75% - 79.43% ) |
|  | LDBio +ve | 28 | 41 | 79.74% | 83.02% |  |
| IgG *Aspergillus fumigatus* |  | Non-CPA | CPA | 75% | 90.35% | 85.06% |
|  | IgG -ve | 103 | 15 | (62.14% - | (83.39% - | (78.88% - 90.00% ) |
|  | IgG +ve | 11 | 45 | 85.28%) | 95.08%) |  |
| Symptoms > 4 weeks and prior history of PTB |  |  |  |  |  |  |
| LDBio LFA |  | Non-CPA | CPA | 75.51% | 76.12% | 75.86% |
|  | LDBio -ve | 51 | 12 | (61.13% - | (64.14% - | (67.04% - 83.32% ) |
|  | LDBio +ve | 16 | 37 | 86.66%) | 85.69% |  |
| IgG *Aspergillus fumigatus* |  | Non-CPA | CPA | 79.59% | 91.04% | 86.21% |
|  | IgG -ve | 61 | 10 | (65.66% - | (81.52% - | (78.57% - 91.91% ) |
|  | IgG +ve | 6 | 39 | 89.76%) | 96.64%) |  |
| Symptoms > 3 months with past history of tuberculosis |  |  |  |  |  |  |
| LDBio LFA |  | Non-CPA | CPA | 75.00% | 74.00% | 74.47% |
|  | LDBio -ve | 37 | 11 | (59.66% - | (59.66% - | (64.43% - 82.91% ) |
|  | LDBio +ve | 13 | 33 | 86.81%) | 85.37%) |  |
| IgG *Aspergillus fumigatus* |  | Non-CPA | CPA | 77.27% | 94.00% | 86.17% |
|  | IgG -ve | 47 | 10 | (62.16% - | (83.45% - | (77.51% - 92.43% ) |
|  | IgG +ve | 3 | 34 | 88.53%) | 98.75% |  |
| Symptoms > 4 weeks with lung cavity |  |  |  |  |  |  |
| LDBio LFA |  | Non-CPA | CPA | 69.23% | 67.74% | 68.67% |
|  | LDBio -ve | 21 | 16 | (54.90% - | (48.63% - | (57.56% - 78.41% ) |
|  | LDBio +ve | 10 | 36 | 81.28% | 83.32%) |  |
| IgG *Aspergillus fumigatus* |  | Non-CPA | CPA | 76.92% | 96.77% | 84.34% |
|  | IgG -ve | 30 | 12 | (63.16% - | (83.30% - | (74.71% - 91.39% ) |
|  | IgG +ve | 1 | 40 | 87.47%) | 99.92%) |  |
| Symptoms > 4 weeks excluding abpa |  |  |  |  |  |  |
| LDBio LFA |  | Non-CPA | CPA | 67.80% | 82.35% | 77.02% |
|  | LDBio -ve | 84 | 19 | (54.36% - | (73.55% - | (69.74% - 83.27% ) |
|  | LDBio +ve | 18 | 40 | 79.38%) | 89.19%) |  |
| IgG *Aspergillus fumigatus* |  | Non-CPA | CPA | 76.27% | 95.10% | 88.20% |
|  | IgG -ve | 97 | 14 | (63.41% - | (88.93% - | (82.19% - 92.74% ) |
|  | IgG +ve | 5 | 45 | 86.38%) | 98.39%) |  |
| Symptoms > 4 weeks with bronchiectasis and excluding abpa |  |  |  |  |  |  |
| LDBio LFA |  | Non-CPA | CPA | 80.56% | 71.79% | 76.00% |
|  | LDBio -ve | 28 | 7 | (63.98% - | (55.13% - | (64.75% - 85.11% ) |
|  | LDBio +ve | 11 | 29 | 91.81%) | 85.00%) |  |
| IgG *Aspergillus fumigatus* |  | Non-CPA | CPA | 80.56% | 94.87% | 88.00% |
|  | IgG -ve | 37 | 7 | (63.98% - | (82.68% - | (78.44% - 94.36% ) |
|  | IgG +ve | 2 | 29 | 91.81%) | 99.37%) |  |

## Discussion

The benefits of LDBio LFA under evaluation reported in existing literature include minimal requirement of resources, time and machinery – all of which are important in diagnosing CPA in resource-constrained settings where CPA is predominantly found. In the present study, in a population presenting with symptoms predominantly suggestive of persistent respiratory symptoms (> four weeks), the assay had a sensitivity, specificity and diagnostic accuracy of 67.6%, 81% and 75.3% respectively. In the population who had a past history of pulmonary tuberculosis, the sensitivity, specificity and diagnostic accuracy of the assay increased to 73.3%, 83.9% and 78.5% respectively. The ImmunoCAP Asp IgG assay for detection of IgG *Aspergillus fumigatus* in the same population had a better sensitivity of ∼ 82.4% and diagnostic accuracy of 82.2% though with similar specificity (82%), using a cutoff of >27 mA/ml. This performance is not as good as published previously at the same cutoff from a previous study.^23^ The possible reason may be the inclusion of serum galactomannan (EIA> 0.5) as microbiological criteria for diagnosis of CPA in the previous study. A significant proportion of controls in our study was also positive for serum galactomannan indicating a plausibility of false positivity owing to previous antibiotics or food habits.^24 25^There was moderate degree of agreement^26^ between LDBio LFA and the ImmunoCAP Asp IgG assay (kappa=0.53).

Diagnosis of CPA is dependent on the use of serological tests especially IgG against *Aspergillus spp* in addition to radiological features^18^. Since the antibody estimation is costly, access to these tests are often restricted in resource constrained settings.^27^ On the other hand, conventional fungal cultures have typically poor positivity rate (ranging from 10-40%) ^28^ which might increase markedly by high volume culture (∼ 54% in mixed cases of pulmonary aspergillosis)^29^. Further, different features on chest radiographs and CT scans have variable sensitivity and specificity which may be as low as ∼28%.^30^ For example, a normal chest radiograph had an excellent performance in ruling out CPA, and such patients do not need testing for *Aspergillus* IgG.^30^ Utility of galactomannan antigen in serum or BAL have been evaluated in numerous studies which have yielded different cut-offs, making it difficult to introduce a uniform criteria for diagnosis of CPA.^31 32 33^Access to point-of-care tests like LDBio LFA, is important in identifying the significant load of CPA patients in tuberculosis-endemic countries which are usually ‘economically developing’ and resource constrained. Our study shows that this assay can be used as a screening test to detect ∼70% of CPA patients. However, the test may be false-negative in ∼ 30% of CPA patients, implying that other ancillary tests have to be performed before ruling out the diagnosis of CPA in those patients with high index of suspicion. Our study also showed that LDBio LFA and ImmunoCAP Asp IgG assay had similar specificity ∼ 82% suggesting that the former may be a reliable ‘rule-in’ test. In the light of the present study, it seems that LDBio LFA, though demonstrating lower sensitivity and comparable specificity IgG *Aspergillus*, may have a significant role in identifying patients with CPA. It can be used a screening test in patients presenting with persistent respiratory symptoms in whom CPA is a probable diagnosis. Due to the low sensitivity of LDBio LFA, those who are negative, need to be followed up with other tests like IgG against *Aspergillus*. Those who are positive, may be treated as cases of CPA, owing to similar specificity of ImmunoCAP Asp IgG assay, provided the clinical and radiological features are compatible with the diagnosis of CPA. An algorithm depicting the possible role of LDBio LFA is depicted in Figure 2.

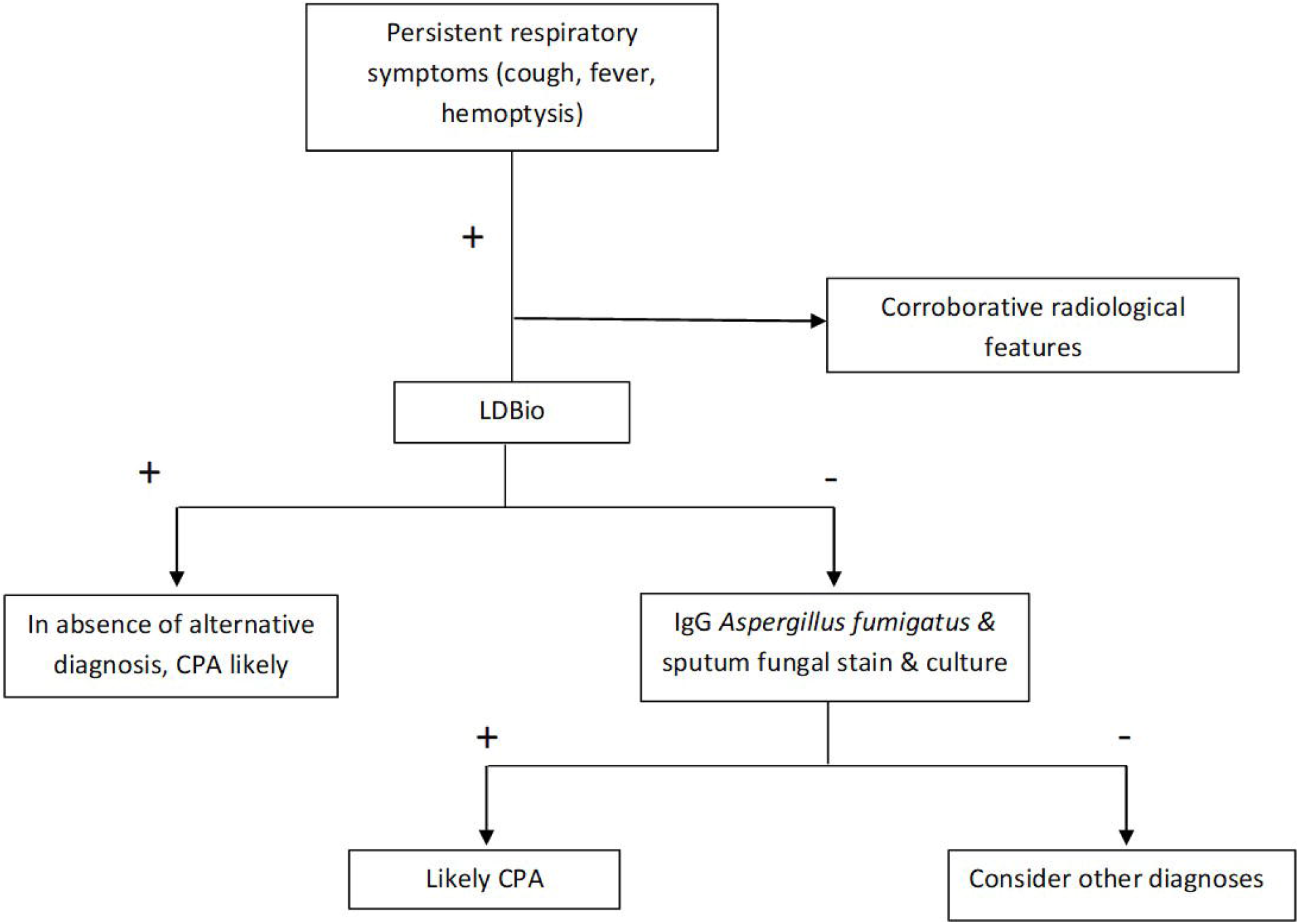

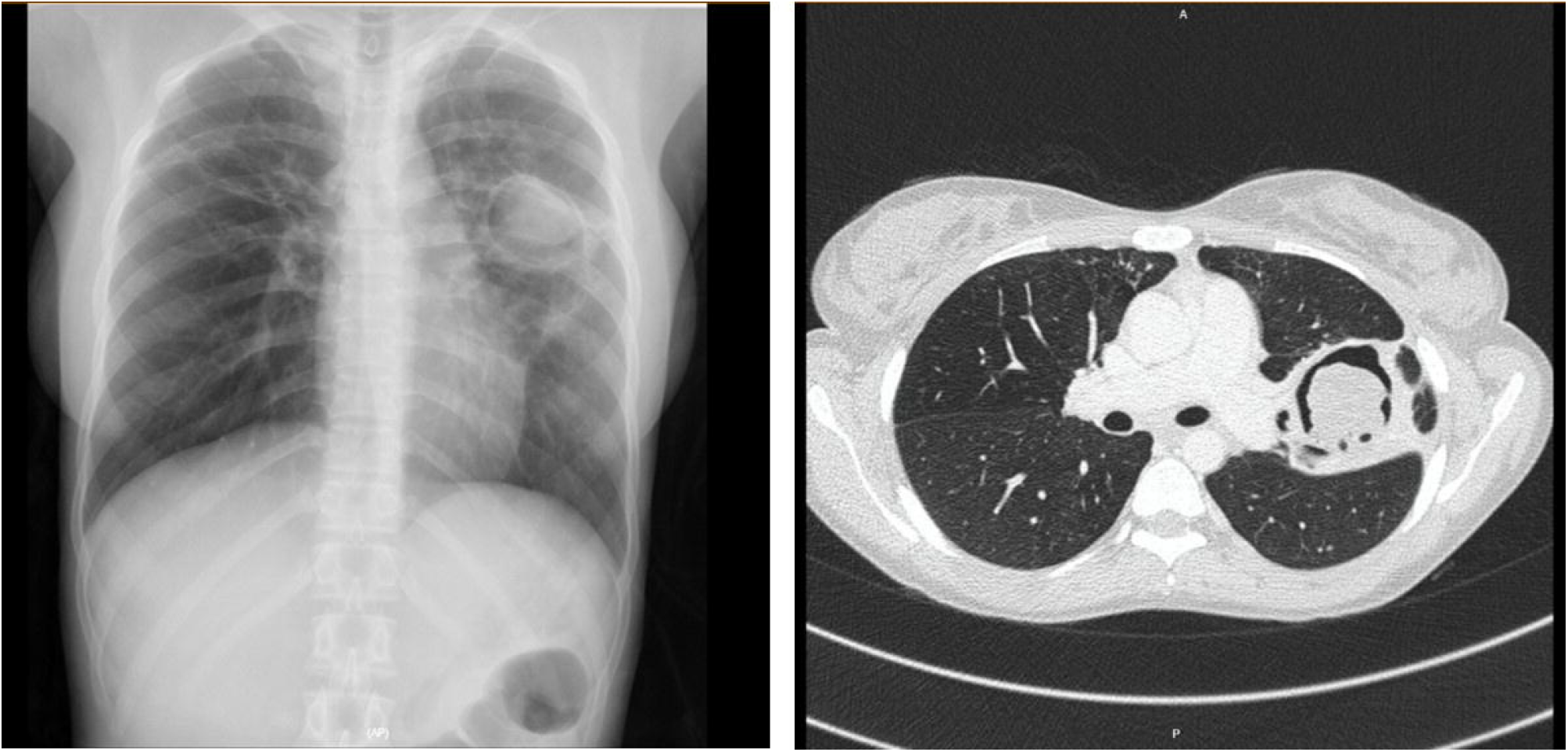

The effectiveness of the LDBio LFA in diagnosing CPA has been demonstrated in three previous studies – one each in France, United Kingdom and Indonesia and its use has also been reported from a case study from Uganda.^34^ The reported sensitivity and specificity in the three studies along with that of our study is shown in table 6. The differences in the sensitivities/specificities of the three studies can be explained by the difference in the recruited population. While, the Indonesian study included patients after completion of tuberculosis therapy, the UK study included sera of known CPA patients and used ‘matched’ sera of healthy controls. In our study, we recruited patients from outpatients and inpatient settings with predominantly chronic respiratory symptoms as the primary presenting complaint. Different diagnostic criteria were used in the various studies. Approximately 66.7% of our patients had past history of pulmonary tuberculosis. Our study population likely represented the real-life scenario where in CPA suspects often present without a past history of respiratory illnesses and often are misdiagnosed as ‘smear-negative’ tuberculosis.^35^ Our study had the following limitations. Only three patients of our patients (<2%) were afflicted by HIV. The absolute number of patients with growth of *Aspergillus* spp in their respiratory samples was also lower (∼ 5.2%) in our study population. However, in our study, all the diagnoses were confirmed using the existing guidelines for diagnosis of CPA and ImmunoCAP Asp IgG, which is widely regarded as a high quality quantitive diagnositic test, was performed in all patients.

| Author | Country/Year | Study type | Population | Comparator | Sensitivity | Specificity | Others |
| --- | --- | --- | --- | --- | --- | --- | --- |
| Piarroux et al<sup>16</sup> | France/2019 | Both retrospective and prospective | All samples received for *Aspergillus* serology(ABPA, CPA, IA SAIA*). Retrospective: 262 cases (68 CPA) & 188 controls. Prospective: 44 cases (11 CPA) & 213 non-cases | Who did not correspond to case definition of CPA | 88.9% | 96.3% | Definition of CPA as per ERS/ESCMID |
| Hunter et al<sup>17</sup> | UK/ 2019 | Cross sectional | CPA patient sera. 154 CPA patients, 150 healthy controls. | Healthy control | 91.6% | 98% | Definition of CPA as per ERS/ESCMID |
| Rozaliyani et al<sup>22</sup> | Indonesia/ 2020 | ? Prospective | Adults with symptoms after completing tuberculosis therapy. | Patients without diagnosis CPA | 80% | 70% | Sputum for fungal culture was used as an essential diagnostic criterion. |
| Ray et al (present study) | India/ 2021-22 present study | Prospective | Patients presenting with respiratory symptoms > 4 weeks. 74 CPA & 100 non-CPA patients | Patients being tested who did not have CPA | 67.6% | 81% | Definition of CPA as per ERS/ESCMID |

## Conclusion

LDBio LFA has reasonable sensitivity and specificity for diagnosis of CPA and can be used in resource-poor settings due to its simplicity of process, minimal requirement for equipment and infrastructure, quick turn-around time and low cost.

## Data Availability

All data produced in the present study are available upon reasonable request to the authors

